# Self-control is associated with health-relevant disparities in buccal DNA-methylation measures of biological aging in older adults

**DOI:** 10.1101/2023.08.30.23294816

**Authors:** Y.E. Willems, A. deSteiguer, P.T. Tanksley, L. Vinnik, D. Främke, A. Okbay, D. Richter, G. G. Wagner, R. Hertwig, P. Koellinger, E.M. Tucker-Drob, K. P. Harden, L. Raffington

**Affiliations:** Max Planck Research Group Biosocial – Biology, Social Disparities, and Development, Max Planck Institute for Human Development, Berlin; Population Research Center, The University of Texas, Austin; School of Business and Economics, Economics Fellow, Tinbergen Institute, Amsterdam; Amsterdam Neuroscience, Complex Trait Genetics, Vrije Universiteit Amsterdam, Amsterdam; Department of Economics, School of Business and Economics, Vrije Universiteit Amsterdam, Amsterdam; Max Planck Institute for Human Development, Berlin; Department of Education and Psychology, Freie Universität Berlin; SHARE Berlin, Berlin; German Socio Economic Panel Study (SOEP), Berlin

**Author notes:** **Corresponding Authors**: Dr. Laurel Raffington, Max Planck Institute for Human Development; Max Planck Research Group Biosocial – Biology, Social Disparities, and Development; Lentzeallee 94, 14195 Berlin, Germany.

**Keywords:** Self-control, DNA-methylation, pace of aging, biological aging, health, life span

## Abstract

Self-control is a personality dimension that is associated with better physical health and a longer lifespan. Here we examined (1) whether self-control is associated with buccal and saliva DNA-methylation (DNAm) measures of biological aging quantified in children, adolescents, and adults, and (2) whether biological aging measured in buccal DNAm is associated with self-reported health. Following preregistered analyses, we computed two DNAm measures of advanced biological age (PhenoAge and GrimAge Acceleration) and a DNAm measure of pace of aging (DunedinPACE) in buccal samples from the German Socioeconomic Panel Study (SOEP-G[ene], *n* = 1058, age range 0-72, *M_age_* = 42.65) and saliva samples from the Texas Twin Project (TTP, *n* = 1327, age range 8-20, *M_age_* = 13.50). We found that lower self-control was associated with advanced biological age in older adults (*β* =-.34), but not young adults, adolescents or children. This association was not accounted for by statistical correction for socioeconomic contexts, BMI, or genetic correlates of low self-control. Moreover, a faster pace of aging and advanced biological age measured in buccal DNAm were associated with worse self-reported health (*β* =.13 to *β* = .19). But, effect sizes were weaker than observations in blood, thus customization of DNAm aging measures to buccal and saliva tissues may be necessary. Our findings are consistent with the hypothesis that self-control is associated with health via pathways that accelerate biological aging in older adults.

## Introduction

Self-control is a dimension of personality that encompasses the ability to delay gratification, inhibit behavioral impulses, and regulate the expression of emotions. Self-control has been proposed to be a key behavioral mediator of both environmental and genetic risk factors for aging-related morbidity and mortality (De Ridder et al., 2018; Duckworth, 2011; Finkenauer et al., 2015; Miller et al., 2015; Moffitt et al., 2011; Robson et al., 2020). Individual differences in self-control arise early in the life course and are associated with myriad health-relevant behaviors and exposures, including sleep, substance use, nutrition, exercise, and socioeconomic attainments (Cobb-Clark et al., 2023; Hoffmann, 2022; Meldrum et al., 2015; Tiemeijer, 2022). These behaviors and exposures have, in turn, been associated with a faster pace of biological aging across multiple physiological systems (Oblak et al., 2021; Pampel et al., 2010; Wang et al., 2022). Little work, however, has directly investigated whether self-control is related to biological aging, which describes the gradual decline in system integrity across tissues and organs that occurs with advancing chronological age (Kirkwood, 2005; López-Otín et al., 2013).

Recently, DNA-methylation (DNAm) measures have been developed to quantify processes of biological aging. DNAm is a stable epigenetic marker that underpins the lifelong maintenance of cellular identity and a dynamic developmental process that changes with age and environmental inputs (Loyfer et al., 2023). Specifically, DNAm measures have been developed to quantify *accelerated biological age* and mortality risk (*e.g.*, GrimAge and PhenoAge Acceleration; Levine et al., 2018; Lu et al., 2019) as well as the *pace of aging* across 18 physiological systems measured repeatedly in the same people (*i.e.,* DunedinPACE, Belsky et al., 2022).

Recent research based on blood samples suggest that lower self-control is associated with accelerated biological age and earlier mortality as indicated by GrimAge Acceleration in 17-50 year old adults (Harvanek et al., 2021; Lei et al., 2022). Moreover, in a five decade prospective study, children with lower self-control later experienced a faster pace of aging in midlife as indicated by analyses of physiological biomarkers (Richmond-Rakerd et al., 2021). As adults, they were also less attentive to practical health information, less consistent in implementing positive health behaviors, and exhibited less positive expectancies about aging. Additionally, those individuals’ self-control in midlife was associated with their pace of aging even after accounting for their self-control in childhood. This suggests that self-control may exert differential influences on aging processes at different points in the life span. It remains unexplored when in the life course associations of self-control with biological aging may become visible; it could take decades until the aging consequences of low self-control arise. DNAm quantifications of biological aging in cohorts of varying ages can help address this question.

While DNAm measures of biological aging are typically developed using blood DNA, buccal and saliva DNA are also commonly collected, particularly in younger cohorts. Buccal and saliva can be sampled via postal kits and this procedure has substantially higher participation rates than blood sampling (e.g., saliva 72% vs. blood 31%, Hansen et al., 2007). Previous findings provide evidence for good saliva-blood cross-tissue correspondence. Blood, saliva and buccal are partially composed of the same cell types: blood samples consist of 100% immune cells, saliva in children consist of approximately ∼35% epithelial cells and ∼65% immune cells (Middleton et al., 2022), and buccal cells in adults consist of ∼80% epithelial cells and ∼20% immune cells (Theda et al., 2018; Wong et al., 2022). While statistical corrections for people’s cell composition are common, immune cell DNAm may be particularly sensitive to early life exposures and aging-related inflammatory processes that can affect multiple tissues, including neurons (Bermick & Schaller, 2022). Additionally, DNAm measures computed in both blood and saliva tissues from the same persons show high cross-tissue rank-order stability (Raffington et al., 2021). More research is needed to assess the applicability of blood-based DNAm measures particularly to buccal tissue, for which cross-tissue rank-order stability appears to be lower than saliva (Raffington et al., 2023).

Here we examined (1) whether self-control is associated with buccal and saliva DNAm measures of biological aging (DunedinPACE, GrimAge Acceleration, and PhenoAge Acceleration) quantified in children, adolescents, and adults, and (2) whether biological aging measured in buccal DNAm is associated with self-reported health. Buccal DNA was collected from participants in the German Socioeconomic Panel Study (SOEP-G[ene], *n* = 1058, age range 0 – 72, *M_age_* = 42.65) and saliva DNA from participants in the Texas Twins Project (TTP, *n* = 1327, 8 – 20, *M_age_* = 13.50). We further tested whether associations differed by chronological age and remained after statistical correction for socioeconomic contexts, body mass index, and smoking, which are commonly associated with DNAm measures of biological aging (Raffington et al., 2021; Raffington & Belsky, 2022), as well as a genetic correlate of low self-control (*i.e.,* a polygenic score of externalizing problems, Karlsson Linnér et al., 2021). We preregistrered our study and highlight where our measures or analyses deviated from our plan (https://osf.io/5sejf, **Supplemental Table 1**). We report standardized beta parameters with 95% confidence intervals and *p*-values corrected for multiple comparisons using Benjamini-Hochberg False-Discovery-Rate (FDR)).

## Results

### (1) Lower self-control is associated with accelerated biological age in buccal tissue from older participants, but not younger adults, adolescents, or children

First, we examined whether self-control was associated with DNAm measures of biological aging. In SOEP-G, we found that lower self-control (as measured by the Brief Tangney Self-control Scale, Tangney et al., 2018) was associated with more advanced PhenoAge and GrimAge Acceleration but not with a faster DunedinPACE (PhenoAge *β* = -.13 [-.25, -.01], *p* = .03; GrimAge *β* =-.15 [-.26, −0.04], *p* =.01; DunedinPACE *β* = -.06 [-0.17, 0.04], *p* = .25). These associations did not survive FDR correction for multiple comparisons. In TTP, children and adolescents’ self-control was not significantly associated with saliva DNAm measures of biological aging (see **Figure 1**, **Supplemental Table 2 and 3**).

**Figure 1.**
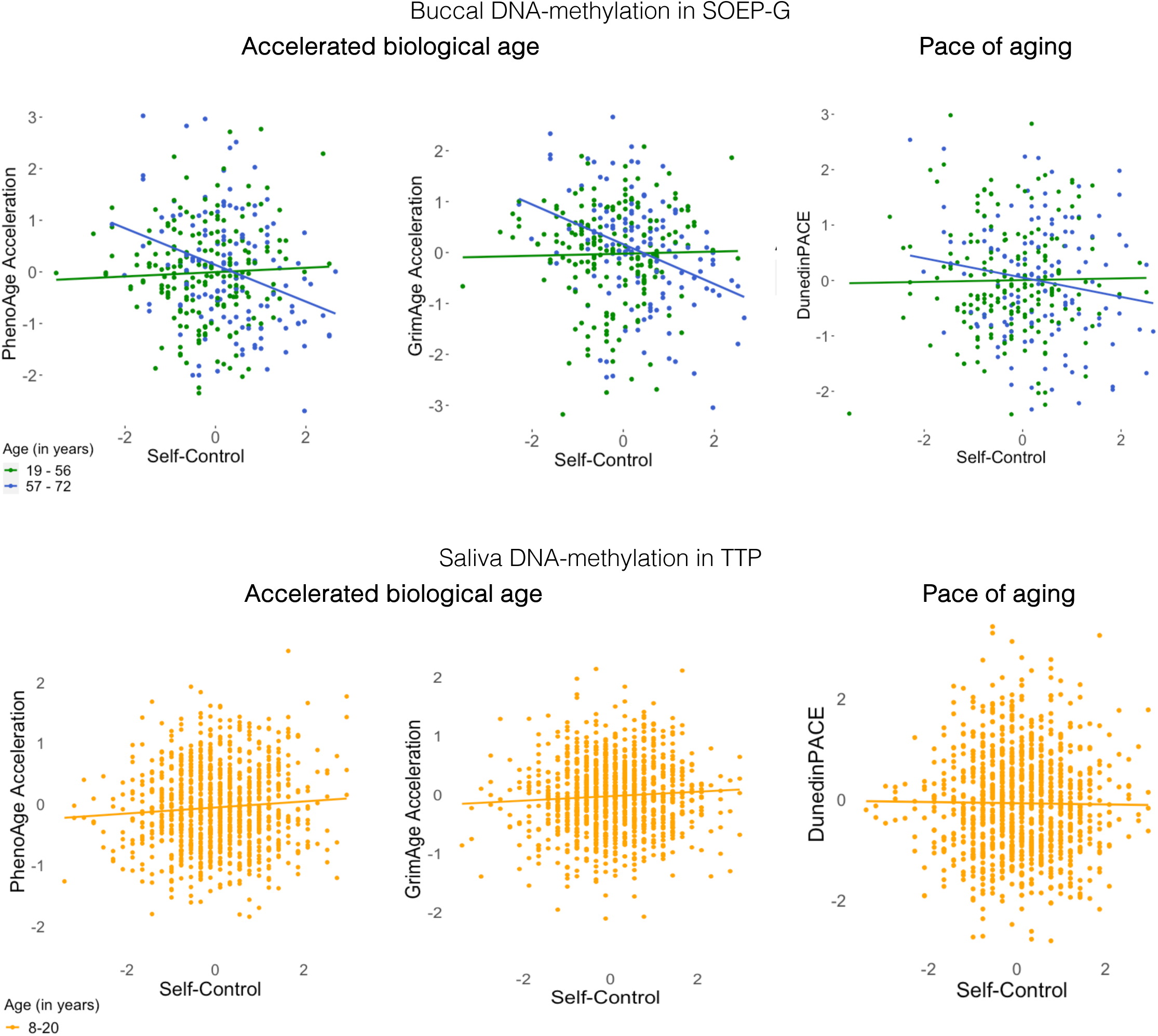
Associations between self-control and DNA-methylation measures of biological aging. *Note:* DNAm-aging measures and self-control are scaled. Self-control was measured with the BTS in SOEP-G and with the grit scale in TTP. See Supplemental Figure 1 for associations of DNAm with attention problems and impulsivity measures in TTP.

Next, we examined whether the association between self-control and DNAm measures of biological aging differed by chronological age in SOEP-G. We found that the association between self-control with PhenoAge and GrimAge Acceleration, but not DunedinPACE, was significantly moderated by chronological age (PhenoAge *β* = -.20 [-.34, -.05], *p* < .01; GrimAge *β* = -.17 [-.28, -.06], *p* <.01; DunedinPace *β* = -.10 [-.24, .03], *p* = .14). These interaction terms remained significant after FDR correction. Accordingly, lower self-control was associated with accelerated biological age in older participants.

To further characterize this age interaction, we stratified participants into older and younger participants by mean split (*M_age_* = 57.02). Among older participants (aged 57-72 years), lower self-control was associated with more advanced PhenoAge and GrimAge Acceleration (PhenoAge *β* = -.34, [-.51, -.17], *p* <.001; GrimAge *β* = -.34, [-.49, -.19], *p* <.001; see **Figure 1**). In contrast, among younger participants (aged 19-56), self-control was not associated with PhenoAge or GrimAge Acceleration (PhenoAge *β* = .06, [-.09, .21], *p* = .45; GrimAge *β* = .03, [-.19, .12], *p* = .66). The association between self-control and DunedinPACE was not statistically significant in younger or older participants (younger *β* =.02 [-0.14, 0.17], *p*=.84; older *β* = -.17, [-.35, .00], *p* = .06; see **Figure 1**).

We have previously found that socioeconomic disadvantage is associated with accelerated buccal PhenoAge and GrimAge and a faster DunedinPACE in SOEP-G (Raffington et al., 2023) as well as a faster saliva DunedinPACE, but not accelerated PhenoAge or GrimAge, in a subsample of TTP children (Raffington et al., 2021). Therefore, we tested whether associations of self-control and DNAm measures of biological aging were accounted for by socioeconomic contexts.

We found that the association of self-control with PhenoAge and GrimAge Acceleration remained statistically significant after controlling for socioeconomic contexts in SOEP-G (see **Supplemental Table 4**). In contrast to a previous analysis of *n*=600 TTP children, which found an association only with DunedinPACE, socioeconomic disadvantage was also associated with accelerated GrimAge in the current sample of *n* =1327 TTP children, even after statistical correction for smoking, BMI, and pubertal timing (*β* =-.13 [-.19, -.07], *p* <.001, **Supplemental Table 5**).

Additionally, associations of self-control with PhenoAge and GrimAge Acceleration in SOEP-G remained statistically significant after controlling for BMI, smoking, and genetic correlates of low self-control (see **Supplemental Table 6** and **7**). Risk aversiveness, which consisted of just 1 response item and was weakly correlated with the Brief-Tangney Self-control scale (*r*=.07, *p*<.05) was not associated with DNAm biological aging measures (see **Supplemental Table 8**). In sum, lower self-control was associated with accelerated biological age in older participants, but not younger adults, adolescents, or children.

### (2) A faster pace of aging and accelerated biological age measured in buccal DNAm are associated with worse self-reported health

Next, non-preregistered analyses evaluated whether buccal DNAm measures of biological aging were associated with self-reported health in SOEP-G. (These analyses focused on SOEP-G as the TTP consists of children and adolescents that are generally in good health).

We found that accelerated biological age and faster pace of aging were significantly associated with more self-reported disease severity (PhenoAge Acceleration: *β* =.13 [.06, .19], *p* <.001; GrimAge Acceleration: *β* =.19 [.12, .26], *p* <.001; DunedinPACE: *β* =.09 [.02, .17], *p*=.01). Accelerated biological age, but not pace of aging, was also associated with worse health as indicated by self-reported general health (See **Figure 2**; PhenoAge Acceleration: *β* =-.12 [-.19, -.05], *p* <.001; GrimAge Acceleration: *β* =-.14 [-.21, -.07], *p* <.001; DunedinPACE: *β* =-.00 [-.08, .07], *p* =.967). There were no significant interaction effects with age (see **Supplemental Table 9**).

**Figure 2.**
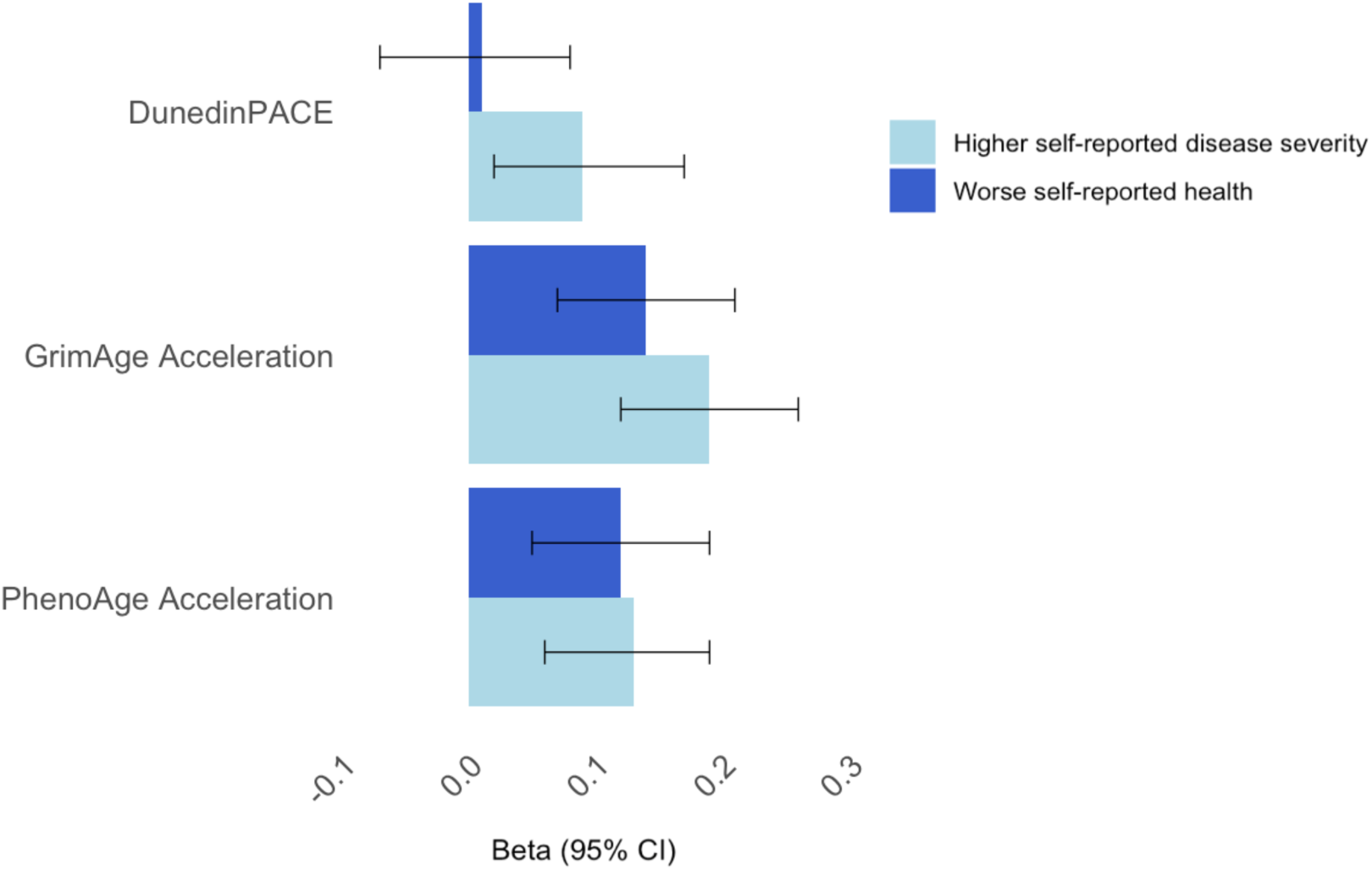
Standardized associations between buccal DNAm measures of biological aging and health in SOEP-G. For illustration purposes, self-reported health is coded such that higher scores reflect worse health.

Next, we tested whether associations of buccal DNAm measures of biological aging with health were statistically accounted for by socioeconomic contexts, BMI, and smoking. We found that the association between DunedinPACE and self-reported disease severity was accounted for by BMI and socioeconomic contexts (see **Supplemental Table 10 and 11**). Associations between PhenoAge and GrimAge Acceleration with self-reported disease severity and health remained statistically significant after accounting for BMI, smoking and socioeconomic contexts (see **Supplemental Table 10 and 11**).

Finally, we examined whether buccal DNAm measures of biological aging statistically accounted for associations of self-control with health. GrimAge Acceleration statistically accounted for 9% of the associations between self-control and self-reported disease severity and health, respectively, in the total sample (indirect effect *β* =-.02, [-.04, -.00], *p*=.03, see **Table 1**). We repeated these analyses for older participants only, for whom self-control was associated with PhenoAge and GrimAge Acceleration (see above). Among older participants, GrimAge Acceleration statistically accounted for 26% of the association between self-control and self-reported disease severity (indirect effect *β* =-.07, [-.14, -.01], *p*=.03, see **Supplemental Table 12**).

**Table 1.** Indirect path estimates of DNA-methylation measures of biological aging statistically accounting for associations of self-control with health.

|  | Accelerated biological age |  |  |  |  |  | Pace of aging |  |  |
| --- | --- | --- | --- | --- | --- | --- | --- | --- | --- |
|  | PhenoAge Acceleration |  |  | GrimAge Acceleration |  |  | DunedinPACE |  |  |
|  | *B* | *95% CI* | *p* | *B* | *95% CI* | *p* | *B* | *95% CI* | *p* |
| *Self-control --> Disease severity* |  |  |  |  |  |  |  |  |  |
| Total Effect | **-.22** | **[-.28, -.16]** | **<.001** | **-.22** | **[-.28, -.16]** | **<.001** | **-.22** | **[-.28, -.16]** | **<.001** |
| Direct Effect | **-.21** | **[-.27, -.15]** | **<.001** | **-.20** | **[-.27, -.14]** | **<.001** | **-.22** | **[-.28, -.15]** | **<.001** |
| Indirect Effects | -.01 | **[-.03, .02]** | .09 | **-.02** | **[-.04, -.00]** | .03 | -.01 | **[-.02, .01]** | .47 |
| *Self-control --> Health* |  |  |  |  |  |  |  |  |  |
| Total Effect | **.22** | **[.15, .29]** | **<.001** | **.22** | **[.15, .29]** | **<.001** | **.32** | **[.15, .29]** | **<.001** |
| Direct Effect | **.21** | **[.14, .28]** | **<.001** | **.21** | **[.14, .27]** | **<.001** | **.32** | **[.15, .29]** | **<.001** |
| Indirect Effects | .01 | **[-.00, .03]** | .10 | **.02** | **[.00, .03]** | .04 | -.00 | **[-.01, .01]** | .85 |

*Note:* Significant associations marked in bold.

## Discussion

We examined (1) whether self-control is associated with buccal and saliva DNAm measures of biological aging quantified in children, adolescents, and adults, and (2) whether biological aging measured in buccal DNAm is associated with self-reported health. First, we found that lower self-control was associated with more advanced biological aging in older participants (57 – 72 years), but not young adults, adolescents or children. The association between self-control with PhenoAge and GrimAge Acceleration in older participants remained statistically significant after controlling for socioeconomic contexts, BMI, smoking, and genetic correlates of self-control. Second, our results indicated that both advanced biological age and a faster pace of aging measured in buccal DNAm were associated with worse self-reported health. While the association between DunedinPACE and self-reported disease severity was accounted for by BMI and socioeconomic contexts, PhenoAge and GrimAge Acceleration were related to self-reported health after accounting for smoking, BMI and socioeconomic status. Thus, despite low cross-tissue correspondence across blood and buccal measures (Raffington et al., 2023), buccal DNAm measures of biological aging appear to capture aging processes relevant to disease and health. But, effect sizes were weaker than observations in blood (GrimAge and health in buccal *β* =.10 - .20 vs in blood *β*=.10 - .50, Faul et al., 2023; Joyce et al., 2021; Lo & Lin, 2022; McCrory et al., 2020) thus customization of DNAm aging measures to buccal tissues may be necessary to maximize their utility.

Collectively, our findings are consistent with the hypothesis that self-control is associated with health via pathways that accelerate biological aging in midlife and older age. Among older participants, GrimAge Acceleration statistically accounted for 26% of the association between self-control and self-reported disease severity and health. Among younger participants, self-control was not associated with biological aging. The effects of self-control-related behaviors on biological aging are likely to accumulate over time, thus, the aging consequences of low self-control may not be visible in the first few decades of life, when people are generally healthy. Moreover, self-control in childhood shows lower rank order stability and may exert independent influences on later life aging compared to self-control in midlife (Richmond-Raker et al., 2021).

We acknowledge limitations. First, our study is based on cross-sectional data and can therefore not make inferences about the direction of the effects between self-control, biological aging, and health. We cannot disentangle whether differences in self-control cause accelerated aging and worse health or, in reverse, worse health causes lower self-control and advanced biological aging. Similarly, age differences in associations between self-control and biological aging could arise from developmental differences or cohort effects related to generational differences (*e.g.,* environmental toxicants, social structures). Second, our findings are likely to be somewhat tissue specific. It is possible, for example, that self-control is associated with the pace of aging in younger samples when DNAm is quantified in blood rather than saliva. In order to take full advantage of buccal and saliva DNA samples, DNAm algorithms developed in these tissues may be needed. Third, our measures of self-control were limited. Future research measuring self-control across informants, ages, and situations is important to tap into the broader range of real-world capacitieis that comprise this umbrella construct.

In conclusion, we find that self-control is associated with buccal DNA-methylation measures of biological aging in midlife and older adulthood in a health-relevant manner. If the cross-sectional findings reported here are found to be causal, then interventions that are successful in increasing self-control might extend the health span (Friese et al., 2017). Alternatively, people’s proximate environments can be manipulated to put less demand on individual self-control behaviors (Reijula & Hertwig, 2022).

## Methods

### Participants

#### SOEP-G

The Socioeconomic Panel (SOEP) is an ongoing population-based, multi-generational survey study. Part sof the SOEP are the “SOEP core” and the “SOEP-Innovation Sample (SOEP-IS), which are two independent random samples of German Households. The SOEP core consists of a broad set of standard survey questions on socioeconomic and sociodemographic background, SOEP-IS supplements this by incorporating data gathered through special questions and experimental modules. In total, SOEP-IS includes 6,576 participants, who were invited to participate in buccal DNA genotyping as part of the “gene subsample” (SOEP-G; Koellinger et al., 2021). In total, there are polygenic indices available for n=2,063 adults (M_age_= 56.13, SD_age_ =18.72, 54% female), with 98% of participants showing high genetic similarity to European reference groups (see Koellinger et al., 2021).

Based on the availability of funds, residual frozen DNA samples of n=1128 of the SOEP-G sample were selected for DNA-methylation analyses. The inclusion criteria were as following: 1) samples from children and adolescents with residual DNA samples holding at least 50ng of DNA, 2) adults with extending age distribution past 18 years, that had at least 250ng of DNA left, had a call rate of at least 0.975, and did not have participating children in the dataset to maximize number of households, 3) match between genetic sex and self-reported sex (see Raffington et al., 2022 for more details). This resulted in the availability of DNA-methylation data for n=1058 participants (M_age_= 42.42, SD_age_ =21.17, 58% female), for whom polygenic scores are also available (see above). The Vrije Universiteit Amsterdam, School of Business and Economics (application number 20181018.1.pkr730) and the IRB of the Max Planck Society (application number 2019_16) granted ethical approval.

#### TTP

The Texas Twin Project (TTP) is an population-representative longitudinal study investigating children and adolescents in the greater metropolitan areas of Austin, Texas (Harden et al., 2013). It has polygenic and DNAm data available for *n*=1327 children and adolescents (M_age_= 13.50, SD_age_ =3.10, 48% females, 34.6% monozygotic twins, 58.9% dizygotic twins). Participants self-identified as White (59.5%), Hispanic/Latinx-only (10.7%), Black/African-American (10.4 %), Asian (8.5%), and Hispanic/Latinx-White (7.8%). The University of Texas Institutional Review board granted ethical approval.

## Measures

Measures are described in Table 2 and include description of the deviation from our preregistration if applicable. Descriptives are presented in Table 3.

**Table 2.**
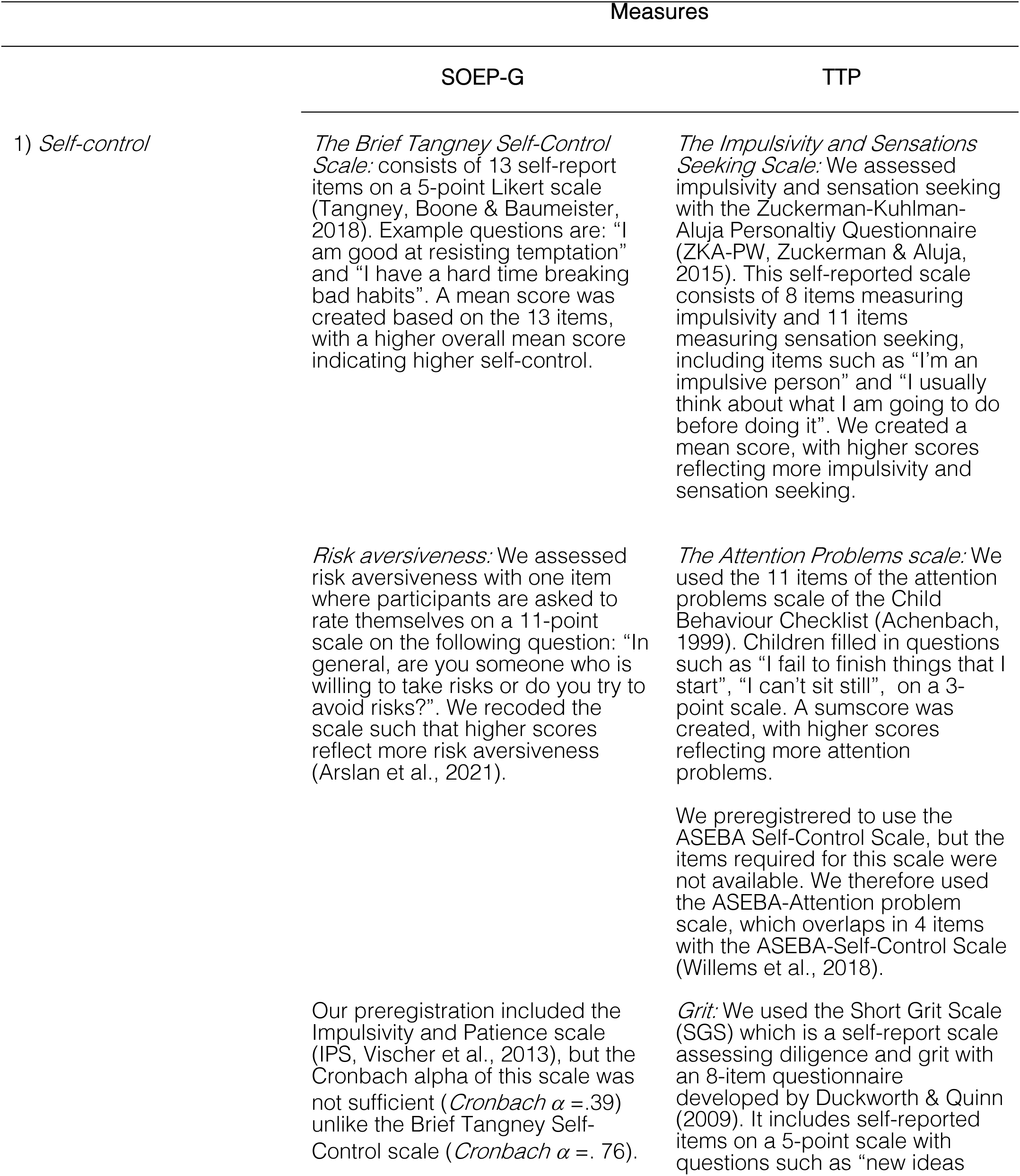
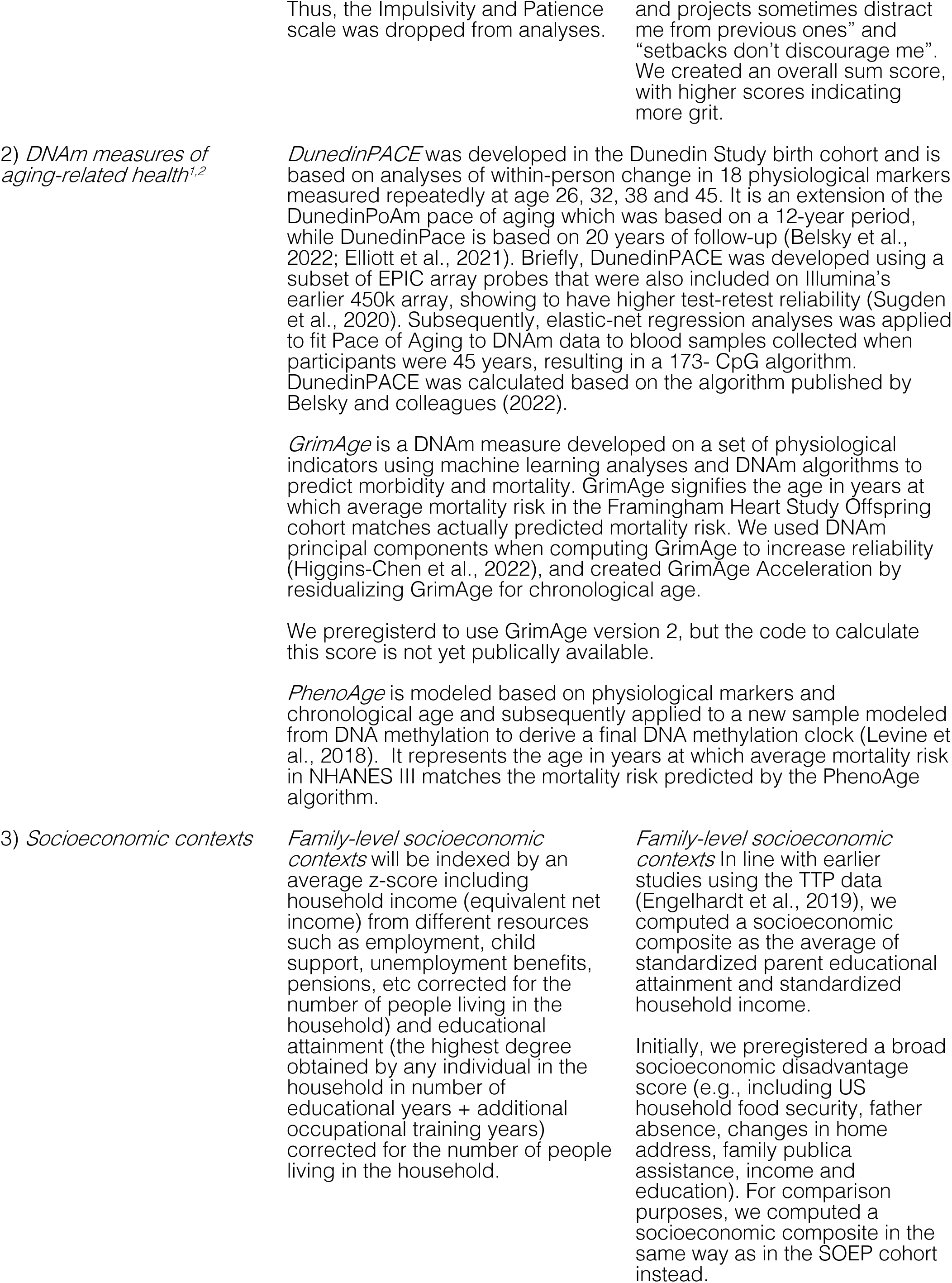
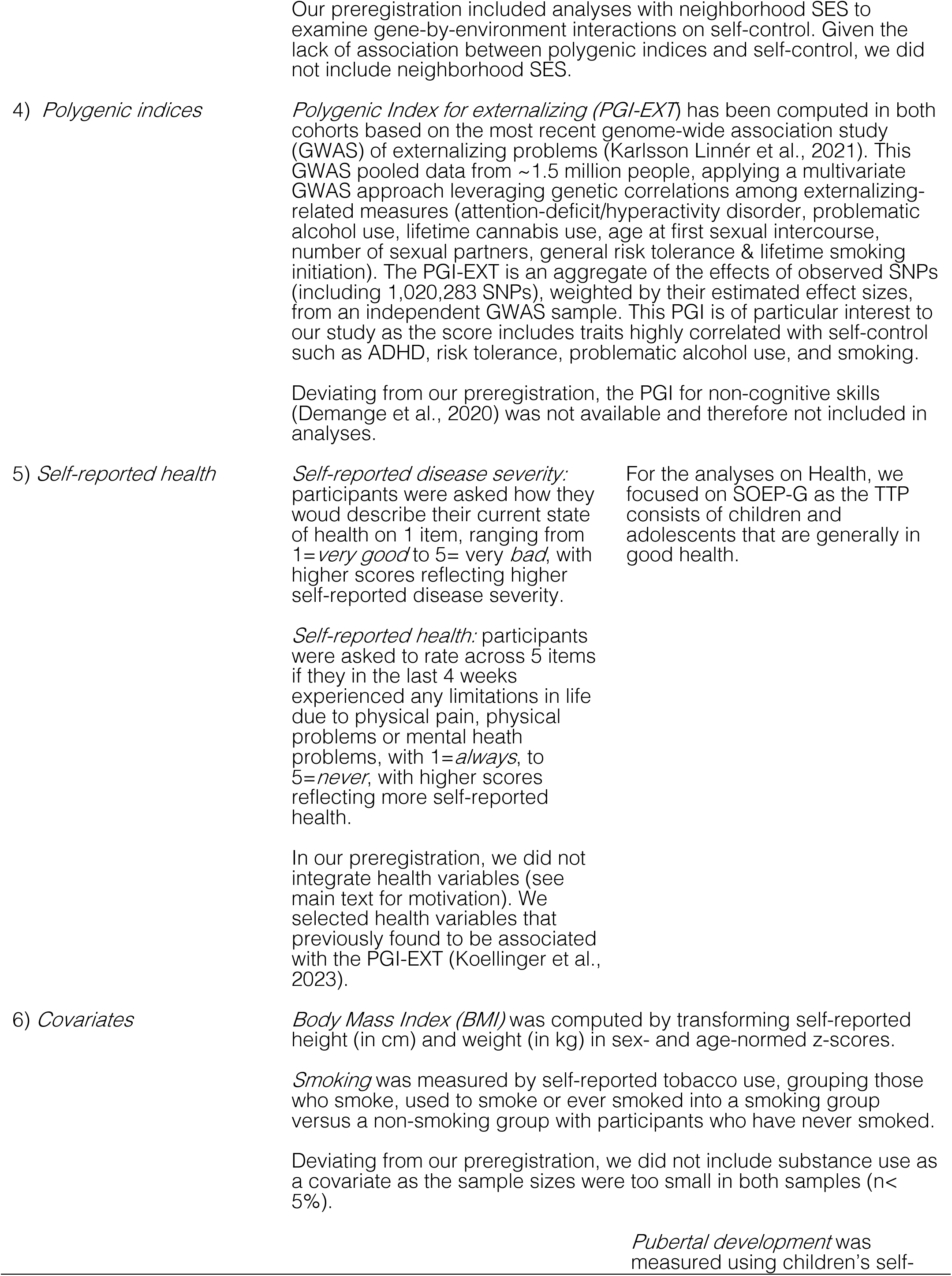
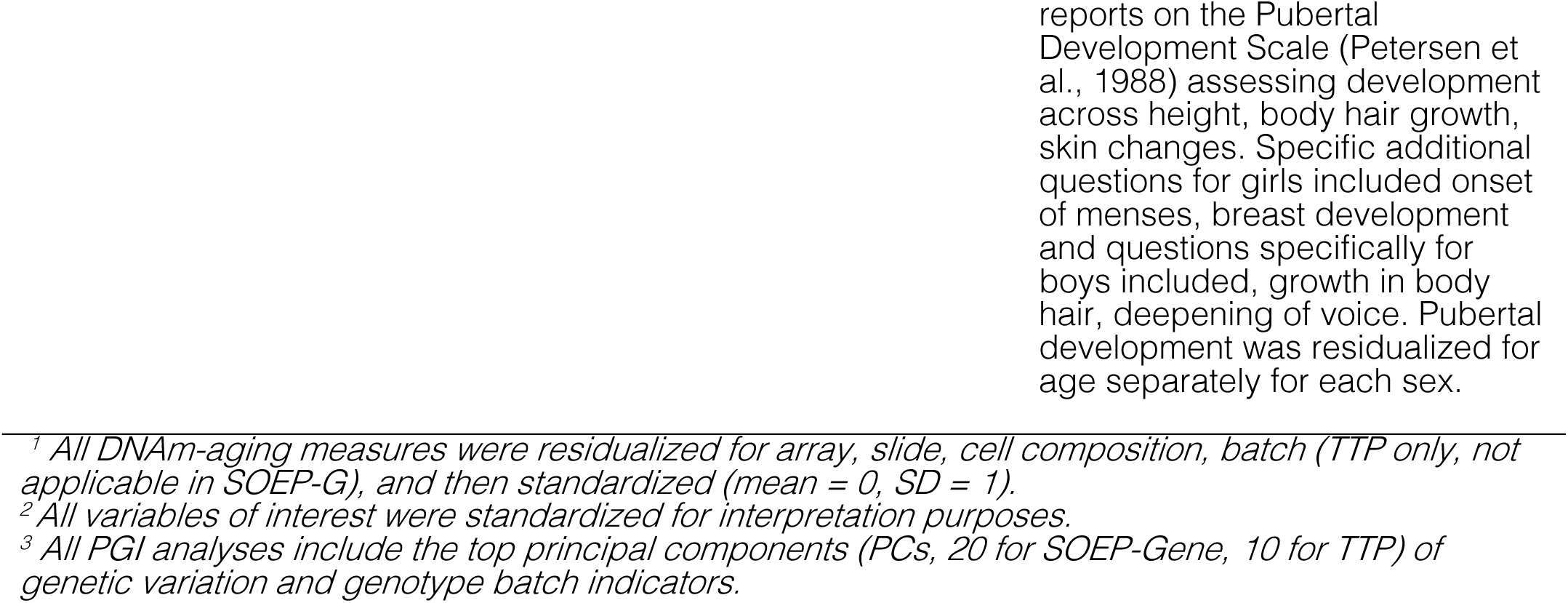
Description of measures.

**Table 3.**
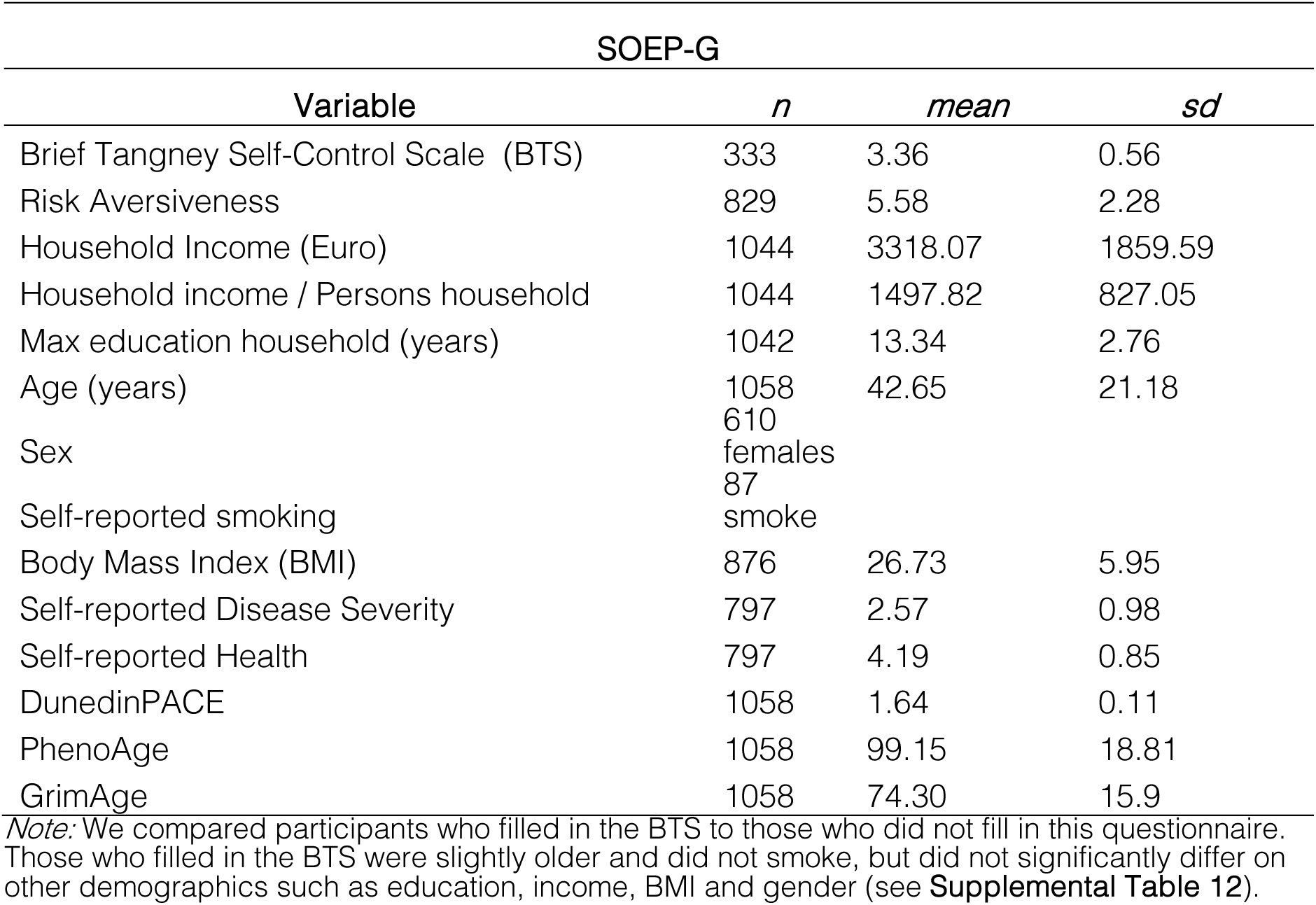
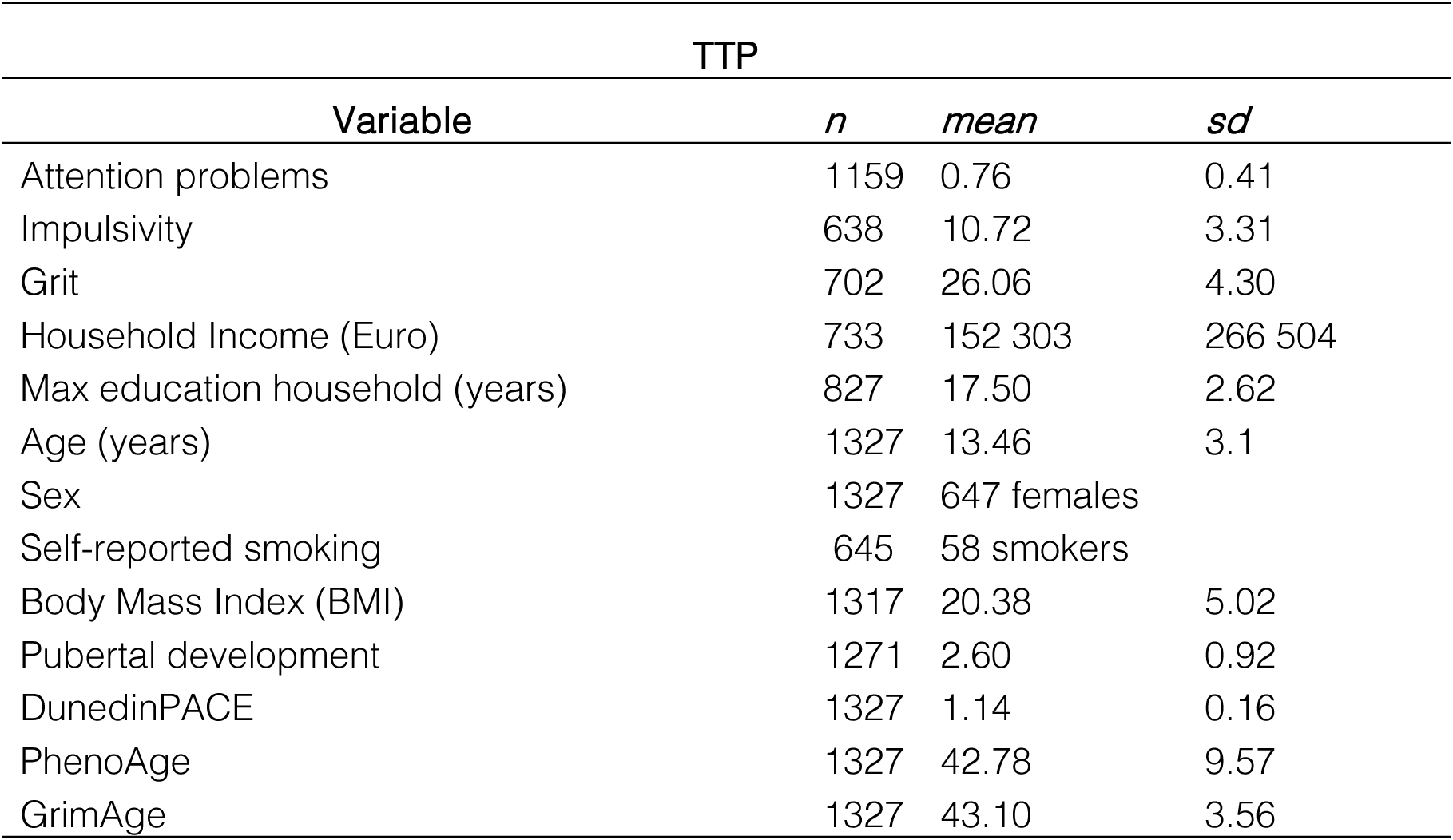
Descriptives for main variables of interest in DNAm subsmaples of SOEP-G and TTP.

### Genotyping

#### SOEP-G

A detailed description of the genetic data in SOEP-G can be found in Koellinger et al., 2023. In short, genotyping was conducted using the Illumina Infinium Global Screening Array-24 v3.0 BeadChips. Genotypes were subject to quality control excluding participants with sex-gender mismatch, with per-chromosome missingness of more than 50%, and with excess heterozygosity/homozygosity.

The Haplotype Reference Consortium reference panel (r.1.1) for imputation was used with imputation accuracy (R2) greater than 0.1. Approximately 66% of the imputed SNPs were rare with minor allele frequencies (MAF) smaller than 0.01 and ∼24% SNPs were common. The average imputation accuracy in the data was 0.66, with higher imputation accuracy for common SNPs (MAF>0.05) with an average imputation accuracy of 0.92. To control for population stratification, the first 20 principal components (PCs) were computed for individuals with high genetic similarity to European reference groups, based on ∼160,000 approximately independent SNPs with imputation accuracy ≥70% and MAF≥0.01 (Koellinger et al., 2021).

#### TTP

The DNA samples were genotyped using the Illumina Infinium PsychArray at the University of Edinburgh, which assays ∼590,000 single nucleotide polymorphisms (SNPs), insertions-deletions (indels), copy number variants (CNVs), structural variants, and germline variants across the genome. Genotypes were subjected to quality control. Briefly, samples were excluded when the call rate was <98% and when there was inconsistent reporting between biological and self-reported sex. Variants were excluded if more than 2% of the data was missing. Untyped variants were imputed on the Michigan Imputation Server, with genotypes being phased with Eagle v2.4 and imputed with Minimac4 (v1.5.7), using the 1K Genomes Phase 3 v5 panel as a reference panel (Auton et al., 2015). Thresholds for minor allele frequency (MAF < 1e-3) and Hardy-Weinberg Equilibrium (HWE p-value < 1e-6) were be applied. Imputed genotypes with poor imputation quality (INFO score < .90) were excluded.

### Preprocessing methylation data

#### SOEP-G

##### Data collection

Buccal swabs and Isohelix IS SK-1S Dri-Capsules were used to collect DNA data. DNA extraction and methylation profiling were conducted at the Erasmus Medical Center in the Netherlands by the Human Genomics Facility (HuGe-F).

##### DNA-methylation data

Methylation levels were assessed using the Infinium MethylEPIC v1 manifest B5 kit at 865,918 CpG sites (Illumina, Inc., San Diego, CA). All samples were from the same batch. DNAm preprocessing was conducted using Illumina’s GenomeStudio software and the packages ‘minfi’, ‘ewastools’ and ‘EpiDISH’ in open-source *R* version 4.2.0 (Aryee et al., 2014; Heiss & Just, 2018; Team, 2013; Zheng et al., 2019). Data cleaning took place in three steps.

First, 20 control metrics were generated in GenomeStudio (see BeadArray Controls Reporter Software Guide from Illumina). Samples were flagged and excluded when falling below the Illumina-recommended cutoffs, including 1) all types of poor bisulfite conversion and all types of poor bisulfite conversion background, 2) all types of bisulfite conversion background <0.5, 3) all types of poor specificity, 4) all types of poor hybridization (excluded n=43). Second, unreliable data points were identified resulting from low fluorescence intensities. Probes with only background signal in a high proportion of samples (proportion of samples with detection p-value > 0.01 is > 0.1) and probes with a high proportion of samples with low bead numbers (proportion of samples with bead number < 3 is > 0.1), were removed. Additionally, cross-reactive probes for Epic arrays and probes with SNPs at the CG or single base extension were also removed (Mccartney et al., 2022; Pidsley et al., 2016). Third, we corrected for background noise and color dye bias (with ‘PreprocessNoob’ in minfi, Triche et al., 2013), accounted for probe-type differences (with ‘BMIQ’ in minfi, Teschendorff & Widschwendter, 2012) and estimated cell composition using robust partial correlations (with ‘HEpiDisch’ in EpiDISH). In order to call the sample a ‘buccal sample’ we set a threshold of 0.5 for epithelial cell proportions (Raffington et al., 2022).

#### TTP

Methylation profiling was conducted by Edinburgh Clinical Research Facility, using the Infinium MethylationEPIC BeadChip kit (Illumina, Inc., San Diego, CA) to assess methylation levels at 850,000 methylation sites. Briefly, preprocessing was conducted with the ‘minfi’ package in R version 4.0.4 (Aryee et al., 2014; R Core Team, 2013). Within-array normalization was performed to address array background correction, red/green dye bias, and probe type I/II correction. To correct for background correction and dye-bias equalization, we applied minfi’s “preprocessNoob” (Triche et al., 2013). Data cleaning took place in three steps. CpG probes were excluded if 1) detection p > 0.01, 2) there were fewer than 3 beads in more than 1% of the samples, 3) they were in cross-reactive regions. Samples were excluded if 1) there was mismatch between self-reported and methylation estimated sex, 2) they showed low intensity probes as indicated by the log of average methylation and their detection p was > 0.01 in >10% of their probes. In R we estimated composition of the immune and epithelial cell types in the samples using “BeadSorted.Saliva.EPIC” within “ewastools” in R, and surrogate variable analyses were used to correct for batch effects (3 batches) using the “combat” function in the SVA package.

## Statistical analyses

Analyses were conducted in R version 4.4.2 and Mplus 8.9 statistical software (RStudio Team, 2020; Muthen & Muthen, 2023). To correct for dependency of observations due to clustering in families (SOEP-G for the PGI analyses) and due to repeated measures within individuals and multiple twin pairs within families (in TTP), we used a sandwich estimator to estimate cluster-robust standard errors. All models included age, gender, and an age-by-gender interaction as covariates, and all variables of interest were standardized for interpretation purposes. We controlled for multiple testing using the Benjamini-Hochberg False-Discovery-rate method (FDR, Benjamini & Hochberg, 1995), and nominal p<.05 for follow-up analyses (covariates and mediation models). See **Table 2** and **Supplemental Table 1** for a list of preregistered analyses and measures and deviations if applicable.

## Data Availability

All data produced in the present study are available upon reasonable request to the authors

