## Supplemental material for "Self-control is associated with health-relevant disparities in buccal DNA-methylation measures of biological aging in older adults"

### Supplement

### Supplemental Tables

**Table S1.** List of preregistered analyses (see https://osf.io/5sejf/), deviations, and results if not reported in main text.

| Research question | Preregistered analysis | Deviation from preregistration, if applicable | Result, if not reported in main text |
|---|---|---|---|
| *Are DNAm measures of biological aging associated with socioeconomic contexts?* | Regress DNAm measures of biological aging on socioeconomic contexts | | For SOEP-G, see Raffington et al., (2023)<br><br>For TTP, see Supplemental Table 5. |
| | Regress DNAm measures of biological aging on socioeconomic contexts including an interaction between socioeconomic contexts and age | | For SOEP-G, see Raffington et al., (2023)<br><br>For TTP, there were no significant socioeconomic context*age interaction effects. |
| | Regress DNAm measures of biological aging on socioeconomic contexts, socioeconomic contexts x age interaction, and covariates. | | For SOEP-G, see Raffington et al., (2023)<br><br>For TTP, see Supplemental Table 5. |
| *Is self-control associated with socioeconomic contexts?* | Regress self-control on socioeconomic contexts. | | In SOEP-G and TTP, there were no statistically significant associations between self-control and socioeconomic contexts. |
| | Regress self-control on socioeconomic contexts, and socioeconomic contexts x age interaction. | | For SOEP-G and TTP, there were no statistically significant age interaction effects. |
| | Regress self-control on socioeconomic contexts, socioeconomic contexts x age interaction, and covariates. | | Not examined given lack of main effect. |

| | | | |
|---|---|---|---|
| *Is self-control associated with DNAm measures of biological aging ?* | Regress DNAm measures of biological aging on self-control. | | See manuscript. |
| | Regress DNAm measures of biological aging on self-control, and self-control x age interaction. | | See manuscript. |
| | Regress DNAm measures of biological aging on self-control, self-control x age interaction and covariates. | | See manuscript. |
| | In TTP, we will fit a bivariate twin model of self-control and DNAm measures of biological aging. | Not examined given lack of association. | |
| | In the subsample of the TTP that have repeated DNAm measures (n=440), we will apply a random-intercept panel model to test longitudinal associations between self-control and DNAm measures of biological aging. | Not examined given lack of association. | |
| *Trivariate associations of self-control, DNAm measures of biological aging , and socio-economic contexts* | If both SES and DNAm measures of biological aging are associated with self-control, we will examine the extent to which this is shared or unique variation in a commonality analysis. | Not examined given lack of association between SES and self-control. | |
| | If both SES and self-control are associated with DNAm measures of biological aging , we will regress DNAm measures of biological aging on both self-control and SES. | | See manuscript. |
| | A frequently cited article by Miller and colleagues (2015) reports that self-control moderates the association of socioeconomic context | | This finding was not replicated. |

| | | | |
|---|---|---|---|
| | and DNAm measures of biological aging. We attempted to replicate this finding by regressing DNAm measures of biological aging on self-control, socioeconomic contexts, self-control x socioecononomic contexts interaction. | | |
| *Is DNAm measures of biological aging associated with PGI?* | Regress DNAm measures of biological aging on PGI. | | For SOEP-G and TTP, DNAm measures of biological aging were not statistically significantly associated with PGI. |
| | Regress DNAm measures of biological aging on PGI, and PGI x age interaction. | | For SOEP-G and TTP, DNAm measures of biological aging there were not statistically significant age*PGI interaction effect. |
| | Regress DNAm measures of biological aging on PGI, PGI x age interaction and covariates. | Not examined given lack of association | |
| | In TTP we test whether dizygotic twin differences in PGI predict DNAm measures of biological aging. | Not examined given lack of association | |
| *Is self-control associated with PGI?* | Regress self-control on PGI. | | For SOEP-G and TTP, self-control was not statistically significantly associated with PGI. |
| | Regress self-control on PGI, and PGI x age interaction. | | For SOEP-G and TTP, there was no statistically significant age*PGI interaction effect. |
| | Regress self-control on PGI, PGI x age interaction, and covariates. | Not examined given lack of association. | |

| | | | |
|---|---|---|---|
| | To test whether the association between self-control and PGI differs by socioeconomic context, we will regress self-control on PGI, PGI x age interaction, PGI x socioeconomic context interaction. | Not examined given lack of association. | |
| | In TTP, we test whether dizygotic twin differences in PGI predict self-control. | Not examined given lack of association | |
| *Multivariate associations of self-control, DNAm, and PGI* | | Not examined given lack of associations with PGI | |
| *Are DNAm measures of biological aging associated with self-reported health?* | | To help contextualize our findings that lower self-control was associated with accelerated biological age, non-preregistered analyses evaluated the association between buccal DNAm measures of biological aging with self-reported health in SOEP-G. (We focused on SOEP-G as the TTP consists of children and adolescents that are generally in good health). | See manuscript |
| *Do DNAm measures of biological aging account for the association of self-control with self-reported health?* | | Not preregistered, see above. | See manuscript. |

**Table S2.** Associations of self-control measures with saliva DNAm measures of biological aging measures in TTP

| | Pace of Aging | | | Accelerated biological age | | | | | |
|---|---|---|---|---|---|---|---|---|---|
| | DunedinPACE | | | PhenoAge Acceleration | | | GrimAge Acceleration | | |
| | *Beta* | *95% CI* | *p* | *Beta* | *95% CI* | *p* | *Beta* | *95% CI* | *p* |
| Attention Problems | .04 | [-.14, .08] | .07 | -.02 | [-.01, .02] | .32 | -.02 | [-.00, .01] | .13 |
| Grit | -.04 | [-.01, .02] | .16 | .02 | [-.08, .05] | .27 | .01 | [-.07, .04] | .40 |
| Impulsivity | .01 | [-.10, .07] | .83 | .01 | [-.07, .05] | .74 | .00 | [-.06, .03] | .89 |

**Table S3.** Associations with saliva DNAm measures of biological aging and Self-control*Age interaction in TTP

| | Interaction | *Beta* | *95% CI* | *p* |
|---|---|---|---|---|
| DunedinPACE | Attention Problems * Age | .01 | [-.04, .03] | .11 |
| DunedinPACE | Grit * Age | -.01 | [-.01, .02] | .41 |
| DunedinPACE | Impulsivity * Age | .01 | [-.04, .03] | .60 |
| PhenoAge Acceleration | Attention Problems * Age | .00 | [-.02, .01] | .60 |
| PhenoAge Acceleration | Grit * Age | -.00 | [-.01, .01] | .48 |
| PhenoAge Acceleration | Impulsivity * Age | -.01 | [-.03, .01] | .38 |
| GrimAge Acceleration | Attention Problems * Age | -.00 | [-.02, .01] | .49 |
| GrimAge Acceleration | Grit * Age | .00 | [-.01, .01] | .62 |
| GrimAge Acceleration | Impulsivity * Age | -.01 | [-.01, .01] | .48 |

**Table S4**. Associations of DNAm measures of biological aging with self-control and SES in SOEP-G.

| | | *Beta* | *95% CI* | *p-value* |
|---|---|---|---|---|
| DunedinPace | BTS | -.06 | [-.17, .05] | .26 |
| | SES | -.10 | [-.22, .02] | .10 |
| PhenoAge Accel | **BTS** | **-.13** | **[-.25, -.01]** | **.03** |
| | SES | -.05 | [-.18, .08] | .42 |
| GrimAge Accel | **BTS** | **-.15** | **[-.26, -.03]** | **.03** |
| | SES | -.08 | [-.21, .04] | .60 |

*Note:* These associations are based on the sample with self-control and DNAm data available ($n$=333), in the full DNAm sample ($n$=1058) the association between SES and DNAm is statistically significant (see Raffington et al., 2023). Significant associations in bold.

**Table S5.** Associations Between Socioeconomic disadvantage and saliva DNAm measures of biological aging in TTP

| | Pace of Aging | | | Accelerated biological age | | | | | |
|---|---|---|---|---|---|---|---|---|---|
| | DunedinPACE | | | PhenoAge Acceleration | | | GrimAge Acceleration | | |
| | *Beta* | *95% CI* | *p* | *Beta* | *95% CI* | *p* | *Beta* | *95% CI* | *p* |
| SES | **-.17** | **[-.27, -.07]** | **<.001** | -.05 | [-.10, .01] | .13 | **-.13** | **[-.19, -.07]** | **<.001** |

*Note:* Significant associations in bold. The association between SES and DunedinPACE holds after correcting for covariates. The significant association between SES and DunedinPACE and GrimAge Acceleration holds after statistically correcting for BMI, pubertal status and smoking.

**Table S6.** Associations of DNAm measures of biological aging and self-control in SOEP-G and BMI

| | | *Beta* | *95% CI* | *p-value* |
|---|---|---|---|---|
| DunedinPace | BTS | -.04 | [-.17, .04] | .49 |
| | **BMI** | **.13** | **[.01 , .19]** | **<.01** |
| PhenoAge Accel | **BTS** | **-.12** | **[-.24, -.01]** | **.04** |
| | BMI | .03 | [-.07, .13] | .54 |
| GrimAge Accel | **BTS** | **-.14** | **[-.26, -.03]** | **.01** |
| | BMI | .03 | [-.07, .12] | .60 |

*Note:* There were no smokers in the BTS sample. Significant associations in bold.

**Table S7.** Associations of DNAm measures of biological aging and self-control in SOEP-G and PGI-Externalizing (PGI-Ext)

| | | *Beta* | *95% CI* | *p-value* |
|---|---|---|---|---|
| DunedinPace | BTS | -.06 | [-.17, .05] | .30 |
| | PGI-Ext | .03 | [-.06, .12] | .49 |
| PhenoAge Accel | **BTS** | **-.14** | **[-.26, -.03]** | **.01** |
| | PGI-Ext | .05 | [-.06, .17] | .34 |
| GrimAge Accel | **BTS** | **-.15** | **[-.25, -.04]** | **<.01** |
| | PGI-Ext | .10 | [-.01, .21] | .07 |

*Note:* Significant associations in bold.

**Table S8.** Associations of DNAm measures of biological aging and risk-taking

| | | *Beta* | *95% CI* | *p-value* |
|---|---|---|---|---|
| DunedinPACE | Risk taking | .02 | [-.05, .09] | .54 |
| PhenoAge Accel | Risk taking | .03 | [-.04, .10] | .36 |
| GrimAge Accel | Risk taking | -.02 | [-.10, .05] | .53 |

**Table S9.** Associations testing the interaction effect of age*DNAm measures of biological aging on health in SOEP-G

| | *Self-reported Disease* | | | *Self-reported Health* | | |
|---|---|---|---|---|---|---|
| Interaction Age*DNAm | *Beta* | *95% CI* | *p-value* | *Beta* | *95% CI* | *p-value* |
| Age*DunedinPACE | -.00 | [-.11, .10] | .47 | -.03 | [-.13, .07] | .58 |
| Age*PhenoAge Acceleration | .08 | [-.02, .13] | .12 | -.07 | [-.18, .04] | .21 |
| Age*GrimAge Acceleration | .05 | [-.05, .15] | .34 | -.04 | [-.15, .08] | .50 |

Note: These analyses are based on a sample of n=797 participants for whom both DNAm and health data was available ($M_{age}$=50,7 , ranging between 19 – 72.50 year old).

**Table S10.** Associations between DNAm measures of biological aging and self-reported disease and health including BMI and Smoking in SOEP-G

| | *Self-reported Disease* | | | *Self-reported Health* | | |
|---|---|---|---|---|---|---|
| | *Beta* | *95% CI* | *p-value* | *Beta* | *95% CI* | *p-value* |
| DunedinPACE | .06 | [-.01, .14] | .09 | .02 | [-.06, .10] | .60 |
| BMI | **.19** | **[.13, .25]** | **< .001** | **-.14** | **[-.21, -.08]** | **< .001** |
| Smoking | .03 | [-.19, .25] | .80 | -.01 | [-.24, .21] | .90 |
| PhenoAge Acceleration | **.13** | **[.06, .20]** | **< .001** | **-.12** | **[-.19, -.05]** | **<.01** |
| BMI | **.20** | **[.13, .26]** | **< .001** | **-.14** | **[-.20, -.07]** | **< .001** |
| Smoking | .02 | [-.19, .24] | .83 | -.00 | [-.22, .22] | .99 |
| GrimAge Acceleration | **.16** | **[.12, .26]** | **< .001** | **-.11** | **[-.18, -.04]** | **<.01** |
| BMI | **.19** | **[.13, .25]** | **< .001** | **-.14** | **[-.20, -.07]** | **< .001** |
| Smoking | -.01 | [-.22, .21] | .96 | .01 | [-.21, .23] | .92 |

Note: Significant associations in bold.

**Table S11.** Associations between DNAm measures of biological aging and self-reported disease and health including SES in SOEP-G

| | Self-reported Disease | | | Self-reported Health | | |
|---|---|---|---|---|---|---|
| | *Beta* | *95% CI* | *p-value* | *Beta* | *95% CI* | *p-value* |
| DunedinPACE | .06 | [-.01, .14] | .09 | .03 | [-.04, .10] | .43 |
| SES | **-.28** | **[-.36, -.21]** | **<.001** | **.29** | **[.21, .36]** | **<.001** |
| PhenoAge Acceleration | **.10** | **[.03, .16]** | **.003** | **-.09** | **[-.16, -.02]** | **.01** |
| SES | **-.28** | **[-.35, -.20]** | **<.001** | **.27** | **[.20, .35]** | **<.001** |
| GrimAge Acceleration | **.15** | **[.08, .22]** | **<.001** | **-.10** | **[-.18, -.03]** | **.01** |
| SES | **-.23** | **[-.29, -.17]** | **<.001** | **.232** | **[.17, .30]** | **<.001** |

Note: Significant associations in bold.

**Table S12.** Indirect path estimates of DNA-methylation measures of biological aging statistically accounting for associations of self-control with health.

| | Accelerated biological age | | | | | | Pace of aging | | |
|---|---|---|---|---|---|---|---|---|---|
| Older sample | PhenoAge Accelleration | | | GrimAge Acceleration | | | DunedinPACE | | |
| *Self-control --> Self-reported disease severity* | *B* | *95% CI* | *p* | *B* | *95% CI* | *p* | *B* | *95% CI* | *p* |
| Total Effect | **-.27** | **[-.40, -.14]** | **<.001** | **-.27** | **[-.40, .14]** | **<.001** | **-.27** | **[-.40, -.14]** | **<.001** |
| Direct Effect | **-.22** | **[-.35, -.09]** | **<.01** | **-.20** | **[-.33, -.07]** | **<.01** | **-.24** | **[-.37, -.12]** | **<.001** |
| Indirect Effects (through mediator) | -.05 | [-.11, .01] | .11 | **-.07** | **[-.14, -.01]** | **.03** | -.03 | [-.07, .01] | .16 |
| *Self-control --> Self-reported health* | | | | | | | | | |
| Total Effect | **.18** | **[.03, .32]** | **.02** | **.18** | **[.03, .32]** | **.02** | **.18** | **[.03, .32]** | **.02** |
| Direct Effect | .14 | [-.02,.29] | .08 | .14 | [-.01, .29] | .07 | **.16** | **[.01, .31]** | **.04** |
| Indirect Effects (through mediator) | .04 | [-.02, .10] | .22 | .04 | [-.03, .10] | .26 | .02 | [-.02, .05] | .33 |

Note: Significant associations in bold.

**Table S13.** Comparing participant who filled in the Brief Tangney Self-control scale to those who did not at key demographics in SOEP-G

| Variable | non-BTS | BTS | t | p |
|---|---|---|---|---|
| Education | 13.39 | 13.22 | 0.91 | .36 |
| Income | 3377.52 | 3191.14 | 1.46 | .14 |
| **Age** | **39.29** | **49.49** | **8.89** | **<.001** |
| BMI | 26.55 | 27.04 | 1.17 | .24 |
| Sex | 58% women | 58% women | 0.13 | .89 |
| **Smokers** | **87 smokers** | **0 smokers** | **9.94** | **<.001** |

Note: Significant associations in bold.

### Supplemental Figures

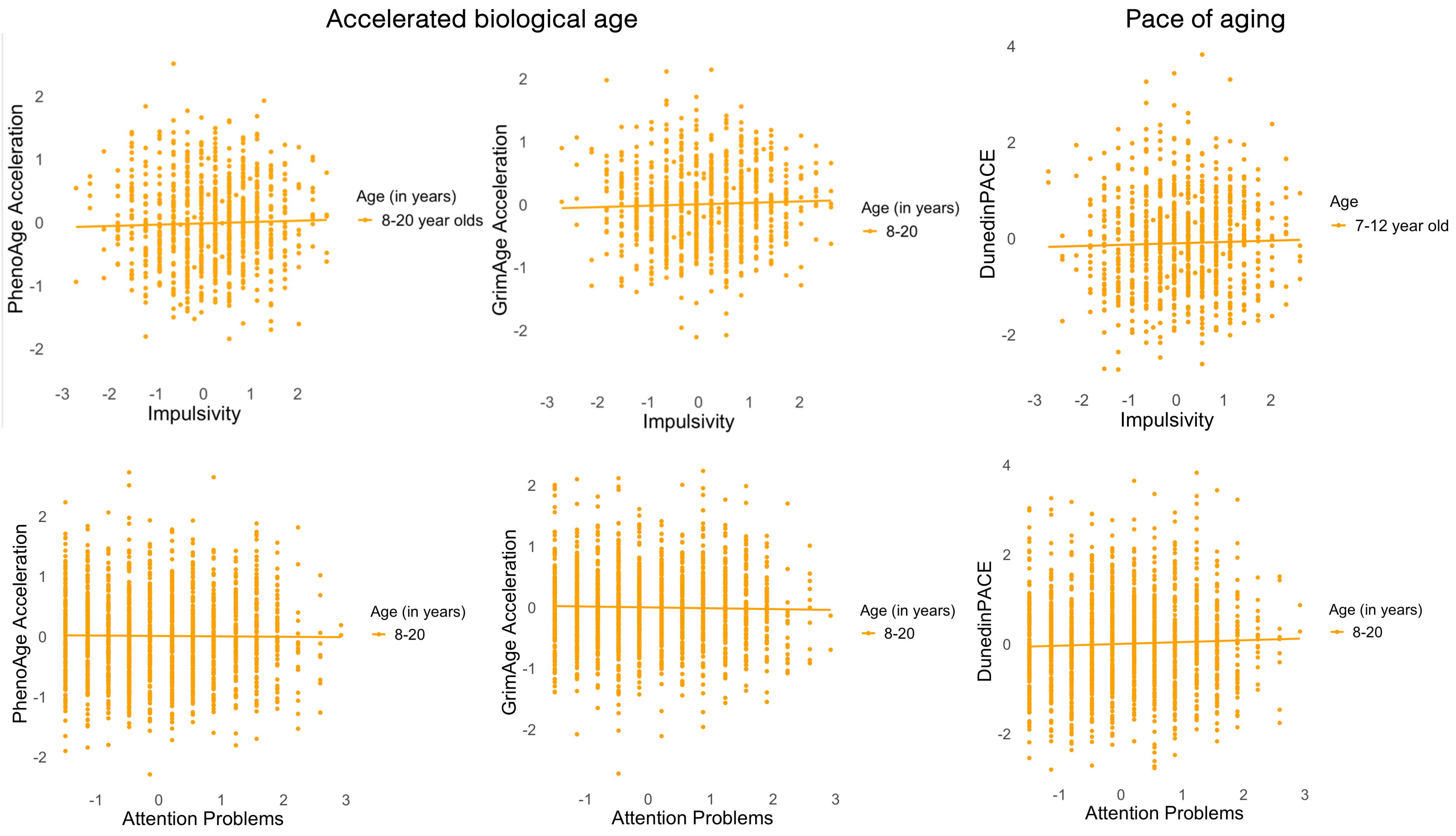

Supplemental Figure 1. Associations between self-control and DNA-methylation measures of biological aging in TTP. DNAm-aging measures and self-control measures are scaled.
